# Epidemiology, risk factors and clinical course of SARS-CoV-2 infected patients in a Swiss university hospital: an observational retrospective study

**DOI:** 10.1101/2020.05.11.20097741

**Authors:** Jean Regina, Matthaios Papadimitriou-Olivgeris, Raphaël Burger, Paraskevas Filippidis, Jonathan Tschopp, Florian Desgranges, Benjamin Viala, Eleftheria Kampouri, Laurence Rochat, David Haefliger, Mhedi Belkoniene, Carlos Fidalgo, Antonios Kritikos, Katia Jaton, Laurence Senn, Pierre-Alexandre Bart, Jean-Luc Pagani, Oriol Manuel, Loïc Lhopitallier

## Abstract

**Background:** Coronavirus disease 2019 (COVID-19) is now a global pandemic with Europe and the USA at its epicenter. Little is known about risk factors for progression to severe disease in Europe. This study aims to describe the epidemiology of COVID-19 patients in a Swiss university hospital.

**Methods:** This retrospective observational study included all adult patients hospitalized with a laboratory confirmed SARS-CoV-2 infection from March 1 to March 25, 2020. We extracted data from electronic health records. The primary outcome was the need to mechanical ventilation at day 14. We used multivariate logistic regression to identify risk factors for mechanical ventilation. Follow-up was of at least 14 days.

**Results:** 200 patients were included, of whom 37 (18·5%) needed mechanical ventilation at 14 days. The median time from symptoms onset to mechanical ventilation was 9·5 days (IQR 7.00, 12.75). Multivariable regression showed increased odds of mechanical ventilation in males (3.26, 1.21-9.8; p=0.025), in patients who presented with a qSOFA score ≥2 (6.02, 2.09-18.82; p=0.001), with bilateral infiltrate (5.75, 1.91-21.06; p=0.004) or with a CRP of 40 mg/l or greater (4.73, 1.51-18.58; p=0.013).

**Conclusions:** This study gives some insight in the epidemiology and clinical course of patients admitted in a European tertiary hospital with SARS-CoV-2 infection. Male sex, high qSOFA score, CRP of 40 mg/l or greater and a bilateral radiological infiltrate could help clinicians identify patients at high risk for mechanical ventilation.

## Introduction

Severe acute respiratory syndrome coronavirus 2 (SARS-CoV-2) first emerged in Wuhan (China) in December 2019. WHO, in February 2020, named the resulting disease COronaVirus Induced Disease 2019 (COVID-19) [1]. There is a wide spectrum of severity ranging from asymptomatic presentation to severe pneumonia requiring ventilator support and death [2], Since March 2020, the pace of SARS-CoV-2 spread around the globe increased as the epidemic evolved to a pandemic [3]. Europe and the USA are now at the epicenter of; the pandemic and face increasing number of severe cases. By April 14 2020, Switzerland registered 25580 cases, 2936 hospitalisation and 885 deaths [4].

Several studies described epidemiological characteristics of COVID-19 patients in China [2,5,6]. To our knowledge, to the exception of a study including critically ill patients [7], none has so far described epidemiology, risk factors and clinical course of COVID-19 patients in a European setting.

We think that it is important to collect and analyse European data. The European population is older, with a likely different genetic background that may influence the evolution of disease and with different resources for responding to the outbreak. Furthermore, although many studies have modelled the risk of severe disease in Chinese patients, little is known; about risk factors for progression to severe disease in Europe [8].

In the present study, we report the epidemiological and clinical characteristics of patients hospitalized for COVID-19 in a Swiss university hospital as well as risk factors for progressive respiratory failure requiring mechanical ventilation.

## Methods

### Study Setting

This study took place in Lausanne University Hospital (LUH), a one-thousand-bed tertiary university hospital in Lausanne, Switzerland. LUH serves as a primary-level community hospital for Lausanne (population circa 300’000 inhabitants) and as a referral hospital for Western Switzerland (population circa 1 – 1.5 million inhabitants). LUH increased its; outbreak response capacity by setting up new intensive care units for the management of COVID-19 patients.

### Study Design and Participants

This retrospective observational study analysed all adult patients consecutively hospitalized with a confirmed SARS-CoV-2 infection from March 1, 2020, to March 25, 2020. Patient’s initial physicians used local guidelines to decide on admission (only patients with risk factors for severe disease or patients needing medical care were hospitalized). For all patients, we ensured of follow-up of up to at least 14 days, or up to discharge or death if they occurred first.

### Data collection

LUH electronic health record (EHR) provided epidemiological, clinical, radiological and laboratory data.

Epidemiological data included age, sex, height, weight, and relevant comorbidities (including a Charlson Comorbidities Index (CCI)). We collected data on clinical presentation, SARS-CoV-2 treatments, concomitant treatments, non-pharmacological interventions and clinical course within LUH.

We recorded radiological findings from reports of chest radiography or computed tomography (CT). We defined healthcare workers as professionals having direct contact with; patients (nurses, physiotherapists, physicians, etc.) or patient samples.

Laboratory data included full blood count, D-dimers, creatinine, highly sensitive cardiac T-troponin, C-reactive protein (CRP), procalcitonin (PCT), ferritin, liver function tests, blood type and real-time PCR to detect SARS-CoV-2 [9].

We entered all data in an electronic clinical report form (eCRF) using the REDCap® platform (Research Electronic Data Capture v8.5.24, Vanderbilt University, Tennessee, USA) [10]. Fellows of the Infectious Diseases, Hospital Preventive Medicine and Internal Medicine Services at LUH entered the data and two of the authors (JR, MP) verified their integrity.

### Clinical management

Treating physicians made all decisions regarding supportive care. Specialists in infectious diseases reviewed all SARS-CoV-2 treatment decisions according to the local recommendations. These included protease inhibitors (ritonavir-boosted lopinavir or atazanavir), hydroxychloroquine or remdesivir. Selected critically ill patients with high inflammatory markers (CRP, D-dimers, PCT and ferritin) received tocilizumab. The choice of treatment depended on drug availability, safety profile drug-drug interactions.

Treating physicians discussed *advance planning of care* and *do not resuscitate orders* with all patients. Limitations of care were decided upon admission according to the patient’s values and goals and the treating physician’s appreciation. We categorized these limitations into two levels: 1) no limitation; 2) limitation to the best supportive care provided in non-monitored wards or intermediate care units but without mechanical ventilation (MV).

### Definition

We defined a confirmed SARS-CoV-2 infection as a positive test for SARS-CoV-2 using real-time polymerase chain reaction (RT-PCR) technology in any respiratory sample.

We classified infection as nosocomial if the onset of symptoms started more than 96 hours after admission for another indication.

We defined obesity either as a body mass index (BMI) of 30 kg/m^2^ or higher, or, when missing anthropometric data, a medical diagnosis of obesity.

We used the Berlin definition for Acute Respiratory Distress Syndrome (ARDS) [11]. We defined *shock* as refractory hypotension requiring infusion of vasopressors. Acute kidney injury (AKI) was identified and classified according to the 2012 Kidney Disease Improving Global Outcome guidelines [12]. We defined liver injury as a 3-fold or greater increase in transaminase levels.

We calculated quick Sequential Organ Failure Assessment (qSOFA) score, Confusion/Respiratory rate/Blood pressure/age > 65 years (CRB-65) score and National Early Warning Score (NEWS) were assessed according to their original descriptions [13–15]. We defined MV as invasive respiratory support through a laryngeal or a tracheostomy tube

### Outcome

The primary outcome was the use of MV for respiratory failure attributed to SARS-CoV-2 pneumonia, within 14 days after admission.

### Statistics

The number of cases diagnosed during the study period defined the sample size. Statistical analyses were performed using R software v3.6.2 (R Foundation for Statistical Computing; www.r-proiect.org). Categorical variables were presented as numbers (percentages), normally distributed continuous variables were presented as mean ± standard deviation (SD) and continuous variables with a skewed distribution were presented as median [interquartile range (IQR)].

For the descriptive analysis, we analysed proportions of categorical variables using chi-square goodness of fit test; we used Student t-test for normally distributed variables or i Mann-Whitney-Wilcoxon test for continuous variables with a skewed distribution.

We used a general linear model based on univariate and multivariate logistic regression to calculate odds ratio for MV. We excluded from the analysis, patients whose care was limited to the best supportive care and patients already mechanically ventilated on admission. To avoid bias resulting from treatment indication, we did not include SARS-CoV-2 specific treatments as variables in the model.

To select predictive variables included in the multivariable analysis model, we first identified clinically relevant variables from the univariate analysis for which the p-value was less than; 0.2. To avoid overfitting, the final multivariate model used only four variables chosen by a backward stepwise approach.

For the inflammatory biomarkers, we converted continuous variables to categorical variables using cut-off values previously identified in the literature [6,16,17]. We did not impute any values for missing data.

### Ethics

This project was conducted in accordance with the Declaration of Helsinki, the principles of Good Clinical Practice and the Swiss Human Research Act (HRA). The project received approval from the Ethics Committee of canton Vaud, Switzerland (2020-00657) that waived the need for informed consent. All data were anonymized before analysis.

## Results

### Epidemiological characteristics

Overall, 200 patients with confirmed SARS-CoV-2 infection were hospitalized at LUH during the study period. In 54 (27.0%), care was agreed to be limited to best supportive care on admission, these patients were older and with more comorbidities (Table 1).

**Table 1.**
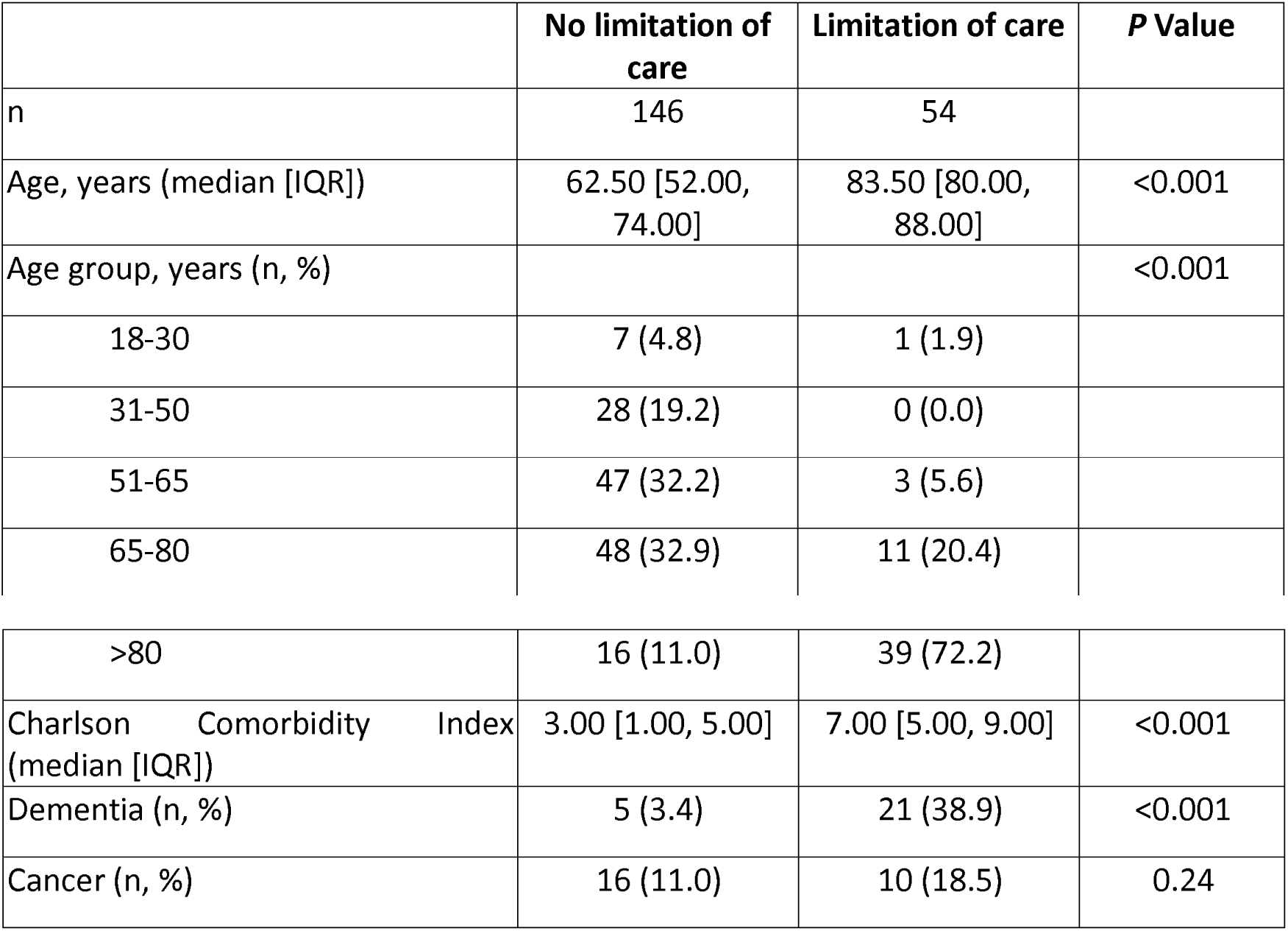
Comparison of patients with and without limitation of care

|  | No limitation of care | Limitation of care | P Value |
| --- | --- | --- | --- |
| n | 146 | 54 |  |
| Age, years (median [IQR]) | 62.50 [52.00, 74.00] | 83.50 [80.00, 88.00] | <0.001 |
| Age group, years (n, %) |  |  | <0.001 |
| 18-30 | 7 (4.8) | 1 (1.9) |  |
| 31-50 | 28 (19.2) | 0 (0.0) |  |
| 51-65 | 47 (32.2) | 3 (5.6) |  |
| 65-80 | 48 (32.9) | 11 (20.4) |  |
| >80 | 16 (11.0) | 39 (72.2) |  |
| Charlson Comorbidity Index (median [IQR]) | 3.00 [1.00, 5.00] | 7.00 [5.00, 9.00] | <0.001 |
| Dementia (n, %) | 5 (3.4) | 21 (38.9) | <0.001 |
| Cancer (n, %) | 16 (11.0) | 10 (18.5) | 0.24 |

Median patient age was 70.0 years [IQR 55.0-81.0], ranging from 20.0 to 96.0 years. Eighty-three (43.0 %) of all inpatients were aged 65.0 years or younger. Median BMI was 26.02 [IQR 22.5-30.71], Overall, 33 (16.5%) patients acquired nosocomial SARS-CoV-2 infection.

One hundred and sixty (80.8 %) patients had at least one comorbidity, with a median Charlson Comorbidity Index of 4.0 [IQR 2.0-6.0]. Table 2 summarizes the characteristics of this population whose most frequent comorbidities were hypertension (43.5%), obesity (27.0%), diabetes (21.5%), coronary artery disease (17.5%) and chronic kidney disease (14.0%).

Fifty-one (26.2 %) patients were treated with angiotensin-converting enzyme inhibitors (ACEI) or angiotensin II receptor blockers (ARBs). Fourteen (7.0 %) were treated with immunosuppressive drugs of which six (3.0 %) patients were transplant recipients (5 solid-organ and 1 haematopoietic stem-cell transplantation). Twenty-six (13.0%) patients had an active malignancy at the time of admission.

**Table 2.**
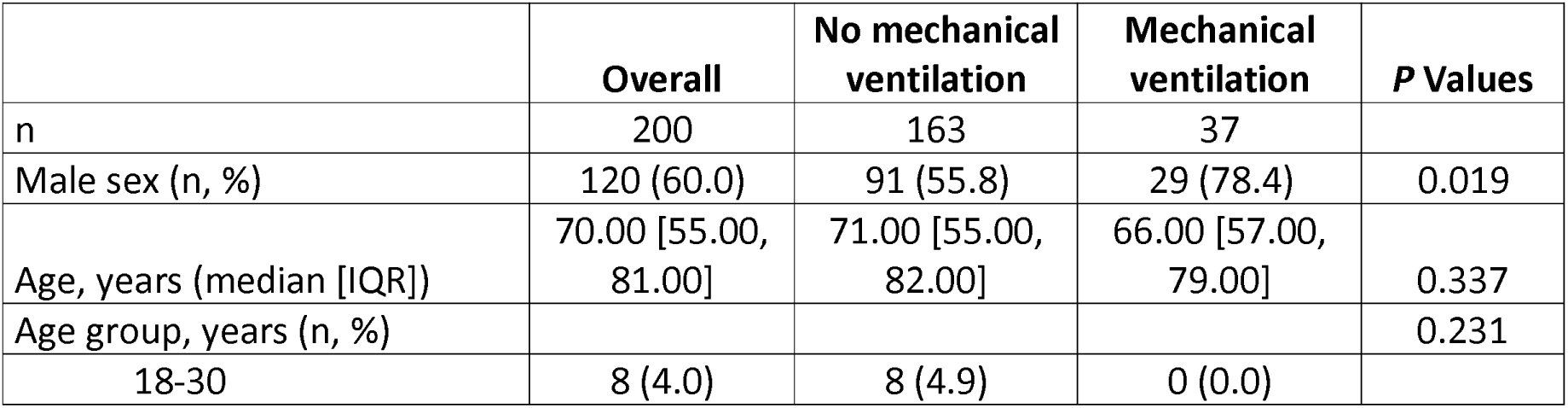
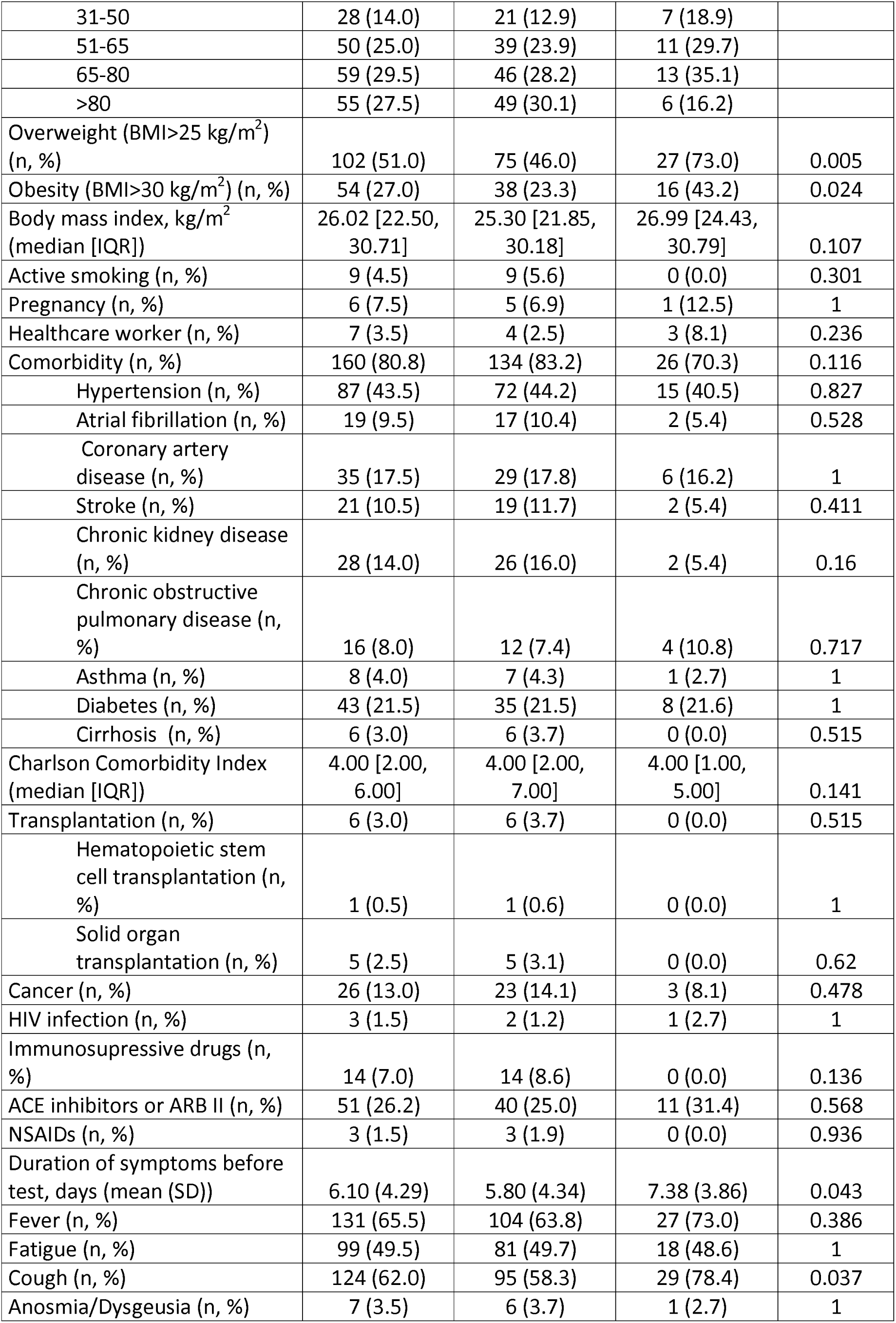
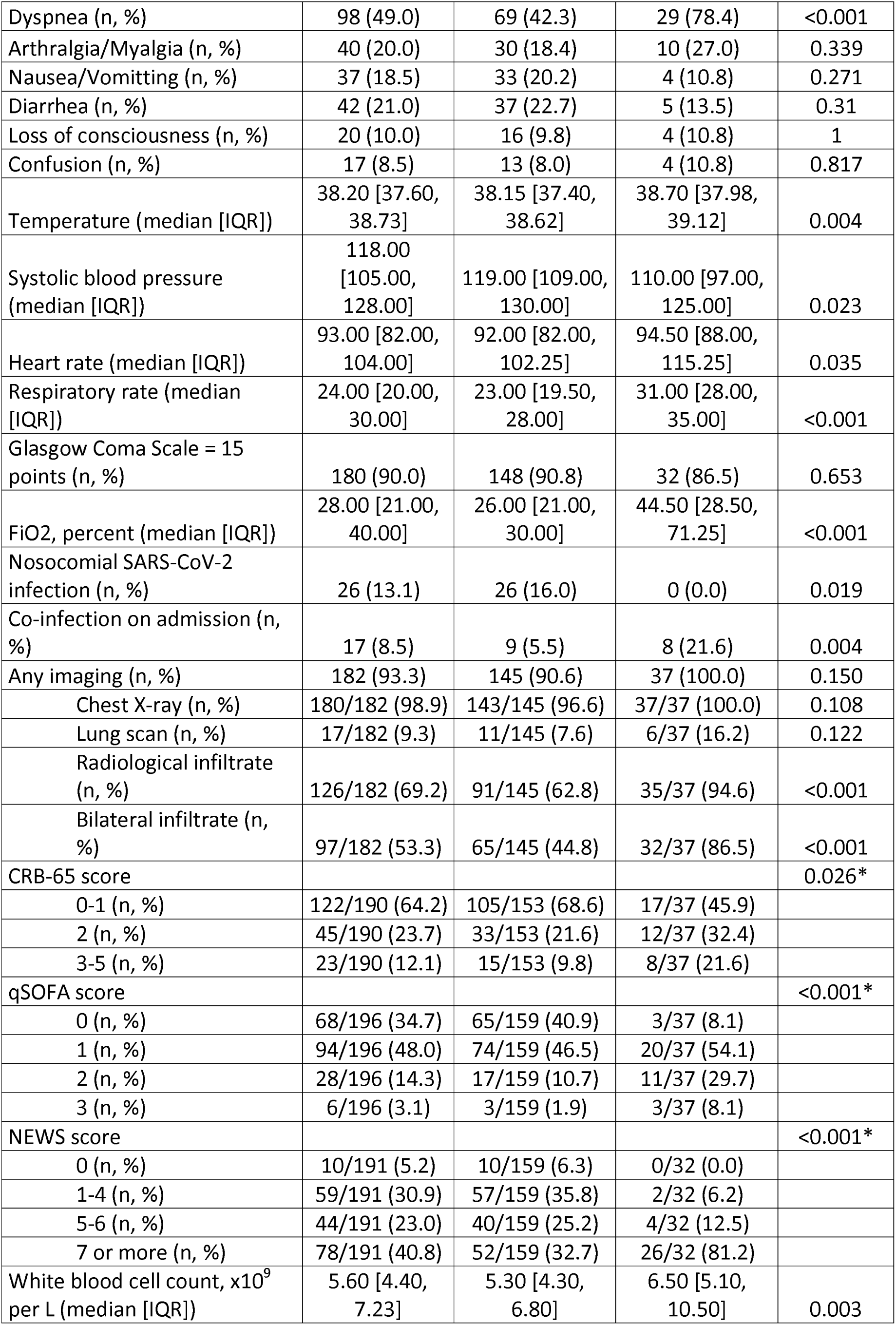
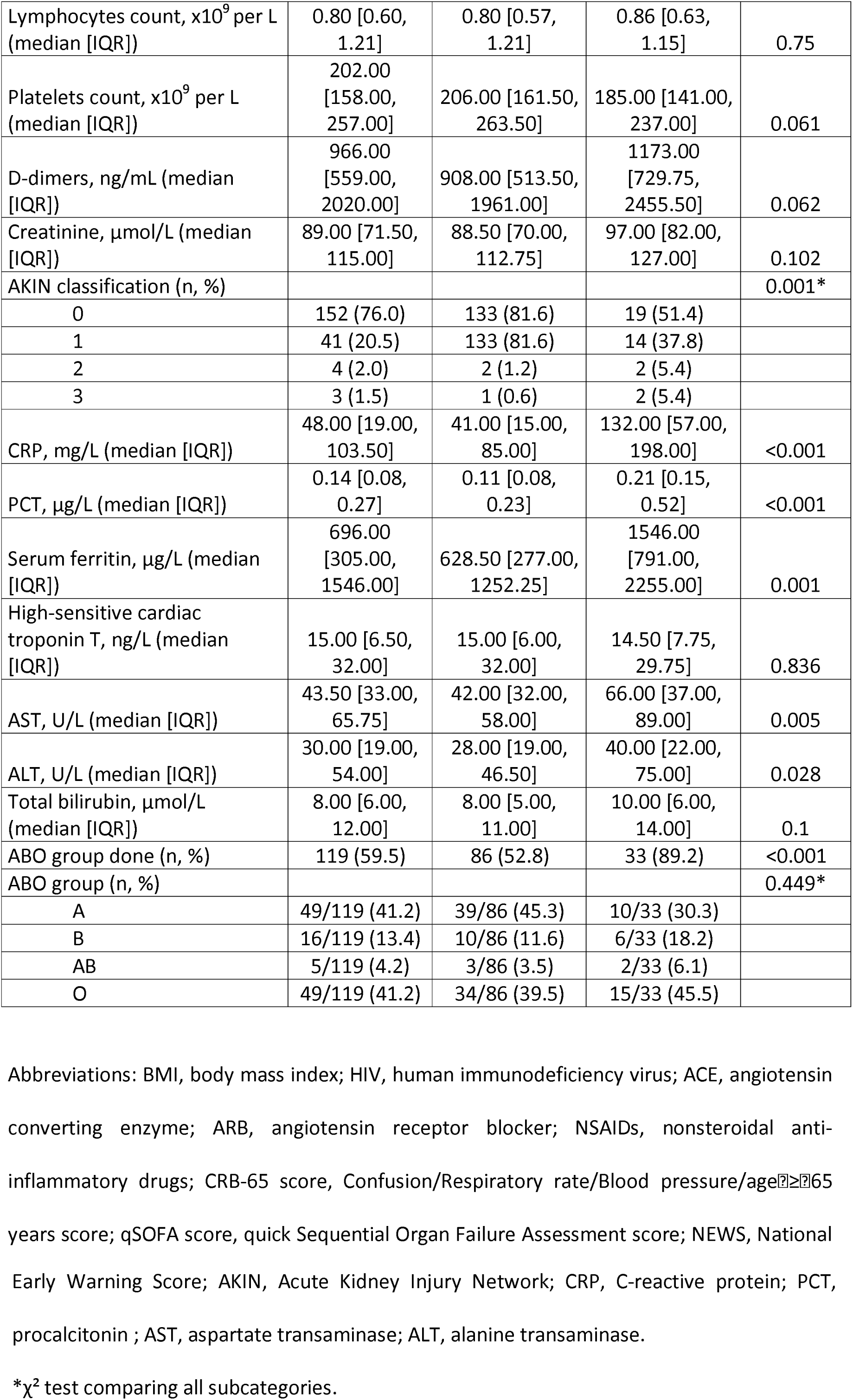
Demographics, clinical, laboratory and radiographic findings of patients on admission

### Clinical characteristics on admission

The mean duration of symptoms preceding admission (or first positive SARS-CoV-2 test for nosocomial cases) was six days (SD 4.29). The most frequent symptoms at the time of testing were fever in 131 patients (65.5%), cough in 124 (62.0%), fatigue in 99 (49.5%) and dyspnoea in 98 (49.0%).

Table 2 describes vital signs, qSOFA score, NEWS and CRB-65 scores on admission.

### Laboratory values and radiology

Table 2 describes the median value of commonly measured inflammatory parameters (white blood cell count, CRP, procalcitonin, D-dimers and ferritin).

Overall, 182 patients (93.3%) had at least one radiological exam, 126/182 of them (69.2%); presented new lung infiltrates which were bilateral for 97/126 (77.0%) of them.

### Treatments

118 (59.0%) of all patients received SARS-CoV-2 treatment (table 3). The most frequently prescribed medication were protease inhibitors in 103 patients (51.5%), hydroxychloroquine in 83 patients (41.5%) and remdesivir in 17 patients (8.5%). Seventeen (8.5%) patients received tocilizumab in adjunction to another SARS-CoV-2 treatment. Seventy-seven patients (38.5%) received two or more concomitant SARS-CoV-2 treatments. Seventy (35.0%) patients received antibiotics. One hundred and thirty-seven (68.5%) patients required supplementary oxygen during the follow-up period.

**Table 3.**
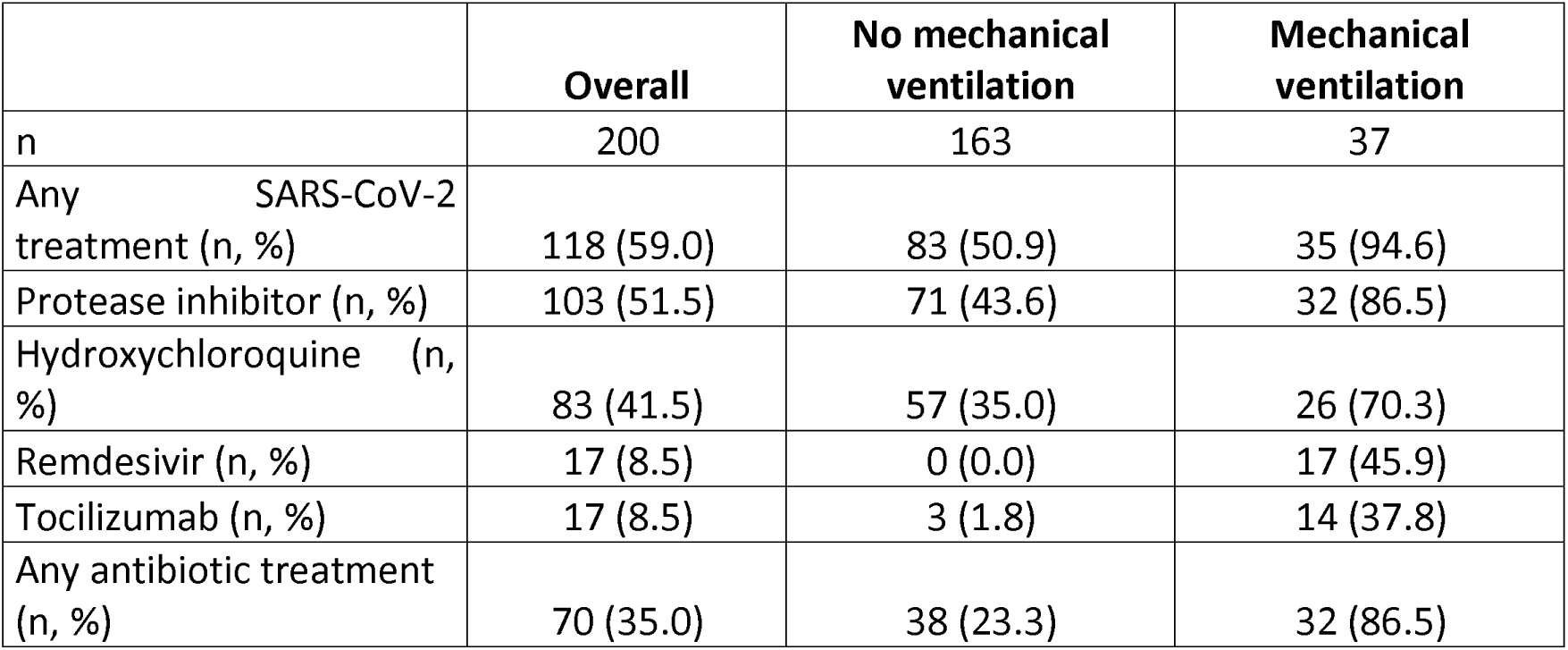
Treatments received by patients during the follow-up period

|  | **Overall** | **No mechanical ventilation** | **Mechanical ventilation** |
| --- | --- | --- | --- |
| n | 200 | 163 | 37 |
| Any SARS-CoV-2 treatment (n, %) | 118 (59.0) | 83 (50.9) | 35 (94.6) |
| Protease inhibitor (n, %) | 103 (51.5) | 71 (43.6) | 32 (86.5) |
| Hydroxychloroquine (n, %) | 83 (41.5) | 57 (35.0) | 26 (70.3) |
| Remdesivir (n, %) | 17 (8.5) | 0 (0.0) | 17 (45.9) |
| Tocilizumab (n, %) | 17 (8.5) | 3 (1.8) | 14 (37.8) |
| Any antibiotic treatment (n, %) | 70 (35.0) | 38 (23.3) | 32 (86.5) |

### Clinical course

Table 4 summarizes patient’s clinical course. At the end of the follow-up, 39 (19.5%) patients were still hospitalized, 101 (50.5%) patients were discharged, 21 (10.5%) patients were transferred to a rehabilitation centre and 14 (7.0%) to another acute care hospital. Twenty-five (12.5%) patients died during hospitalization.

Overall, 38 (19.0%) patients required mechanical ventilation, 37 (97.4%) of them before the 14^th^ day and after a median of 2 days since admission (IQR 0.00 - 3.00). Median time from symptom onset to mechanical ventilation was 9.5 days (IQR 7.00 - 12.75]. Regarding patients requiring MV, 26 (68.4%) had a least one session of prone positioning, 24 (63.2%) received a vasopressor, 11 (28.9%) were eventually weaned from ventilator support and 11 (28.9%) died. The median duration of MV was 6 days (IQR 5.00 - 11.00). Twenty-two of the 38 patients (57.9%) requiring MW were admitted to new dedicated COVID-19 ICUs.

**Table 4.**
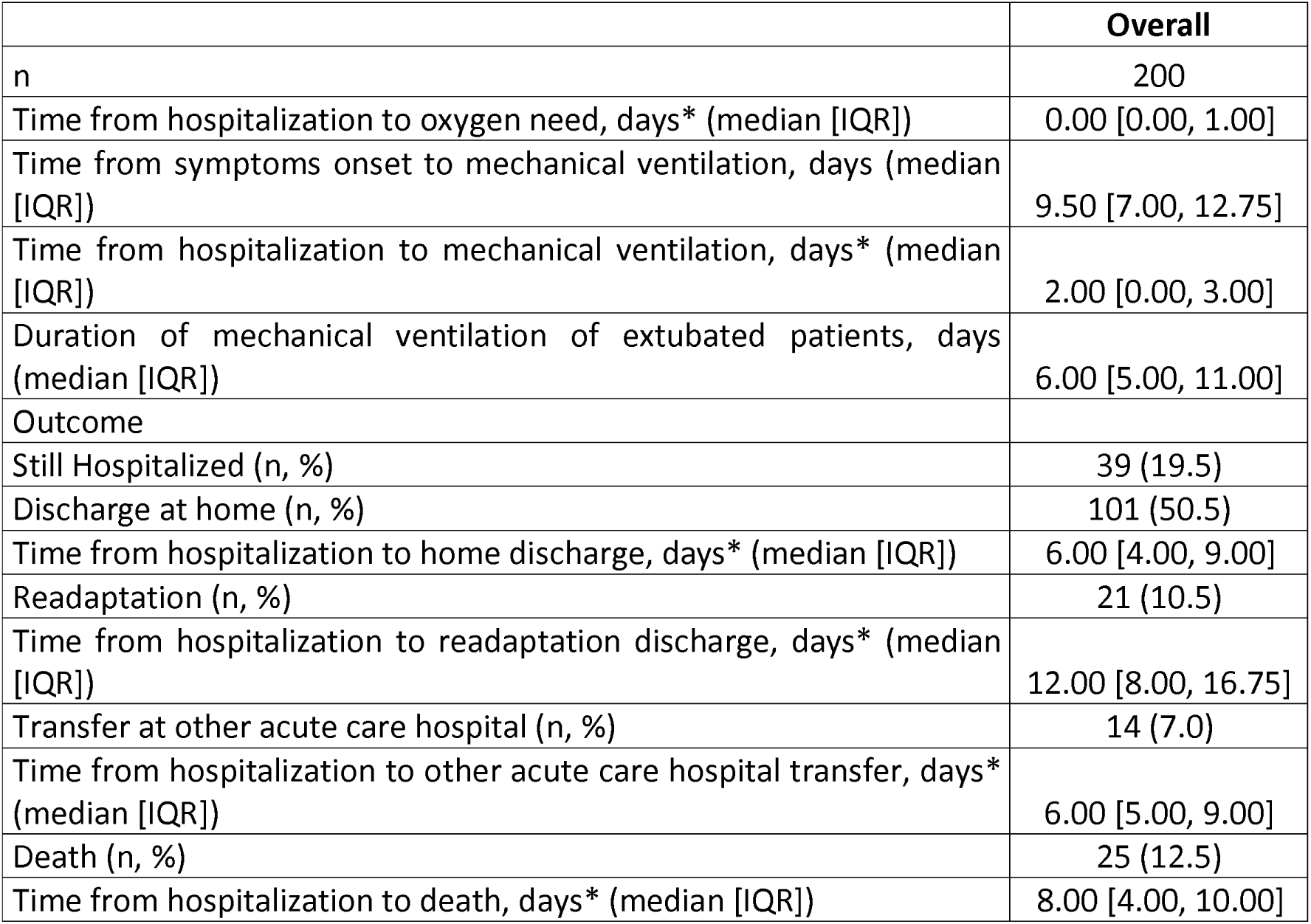
Clinical course

### Complications

Table 5 describes complications during follow-up. The most frequent complications were i ARDS (*n* = 44, 22.0%), acute confusional state (n = 30, 15.0 %), acute kidney injury (*n*=30, 15.0%), hospital-acquired pneumonia (HAP) (*n* = 26, 13.0%) and arrhythmia (*n* = 21, 10.5%) of patients.

**Table 5.**
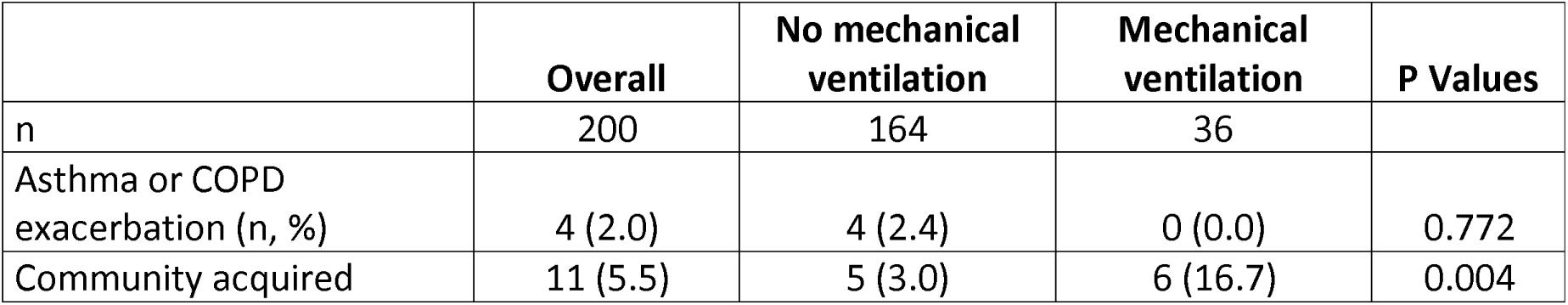
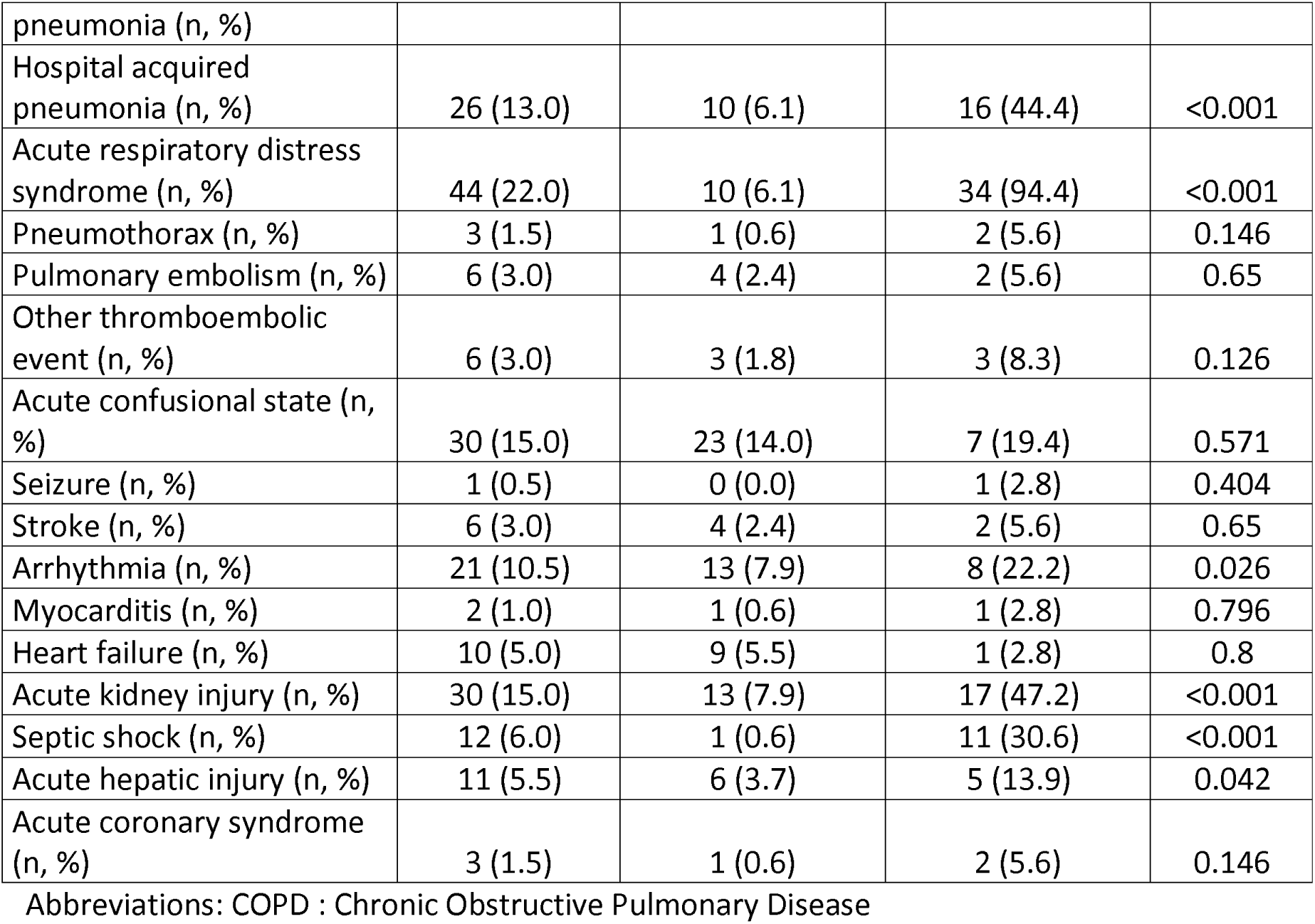
Complications during the follow-up period

### Risk factors for mechanical ventilation

Of 200 patients included in the study, we included 145 (72.5%) in the analyses assessing the risk factors for mechanical ventilation (MV) (figure 1).

**Figure 1.**
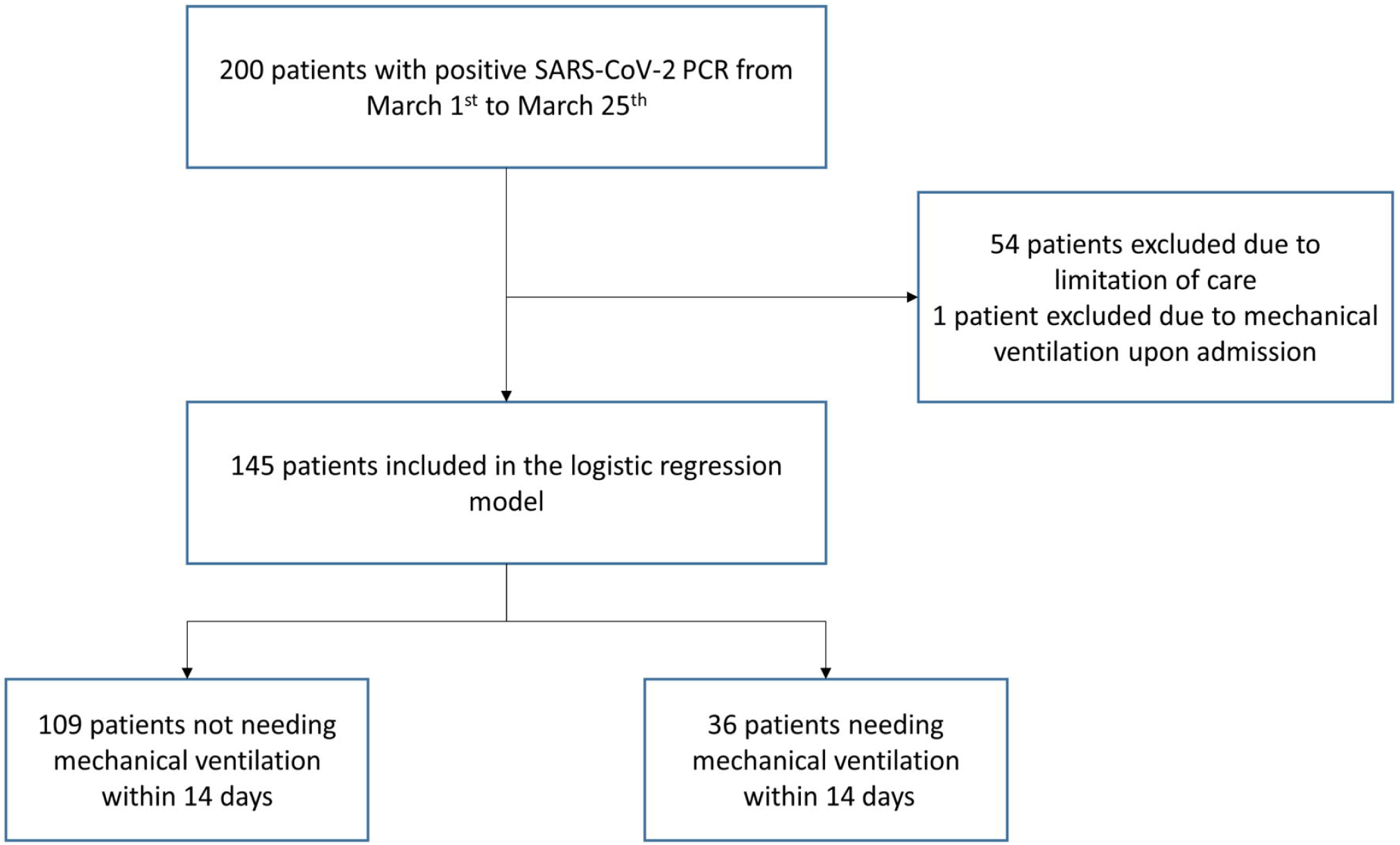
Patient’s selection for univariate and multivariate models.

S1 Table summarizes unadjusted odds of MV at 14 days. Unadjusted odds of MV at 14 days were greater in males (odds ratio 2.65, 95% confidence interval 1.15 to 6.72) and in overweight patients (odds ratio 2.46, 95% confidence interval 1.11 to 5.81). None of the comorbidities increased the unadjusted risk MV at 14 days. Unadjusted odds of MV at 14 days were greater for patients presenting with dyspnoea (odds ratio 4.29, 95% confidence interval 1.86 to 10.85) on admission.

NEWS score ≥7 (odds ratio 9.18, 95% confidence interval 3.66 to 26.55), qSOFA score ≥2 (odds ratio 6.86, 95% confidence interval 2.68 to 18.47) or a CRB-65 score ≥2 (odds ratio 4.64, 95% confidence interval 2.07 to 10.65) increased the unadjusted odds of MV.

The presence of a radiological infiltrates increased the odds of mechanical ventilation (odds ratio 11.0, 95% confidence interval 3.14 to 70.45) as did the presence of a bilateral infiltrates (odds ratio 7.05, 95% confidence interval 2.75 to 21.87). Acute kidney injury on admission (odds ratio 4.27, 95% confidence interval 1.87 to 9.91), D-dimers of 1000 ng/ml or greater (odds ratio 3.28, 95% confidence interval 1.37 to 8.25), CRP of 40 mg/l or greater (odds ratio 6.79, 95% confidence interval 1.51 to 18.58) and PCT of 0.5 μg/l or greater (odds ratio 5.99, 95% confidence interval 1.52 to 29.66) increased the unadjusted odds of mechanical ventilation.

Adjusted odds of mechanical ventilation at 14 days were greater in males (odds ratio 3.26, 95% confidence interval 1.21 to 9.8), in patients who presented with a qSOFA score ≥2 (odds ratio 6.02, 95% confidence interval 2.09 to 18.82), with bilateral infiltrate (odds ratio 5.75, 95% confidence interval 1.91 to 21.06) or with a CRP of 40 mg/l or greater (odds ratio 4.73, 95% confidence interval 1.51 to 18.58) on admission (Table 6).

**Table 6.**
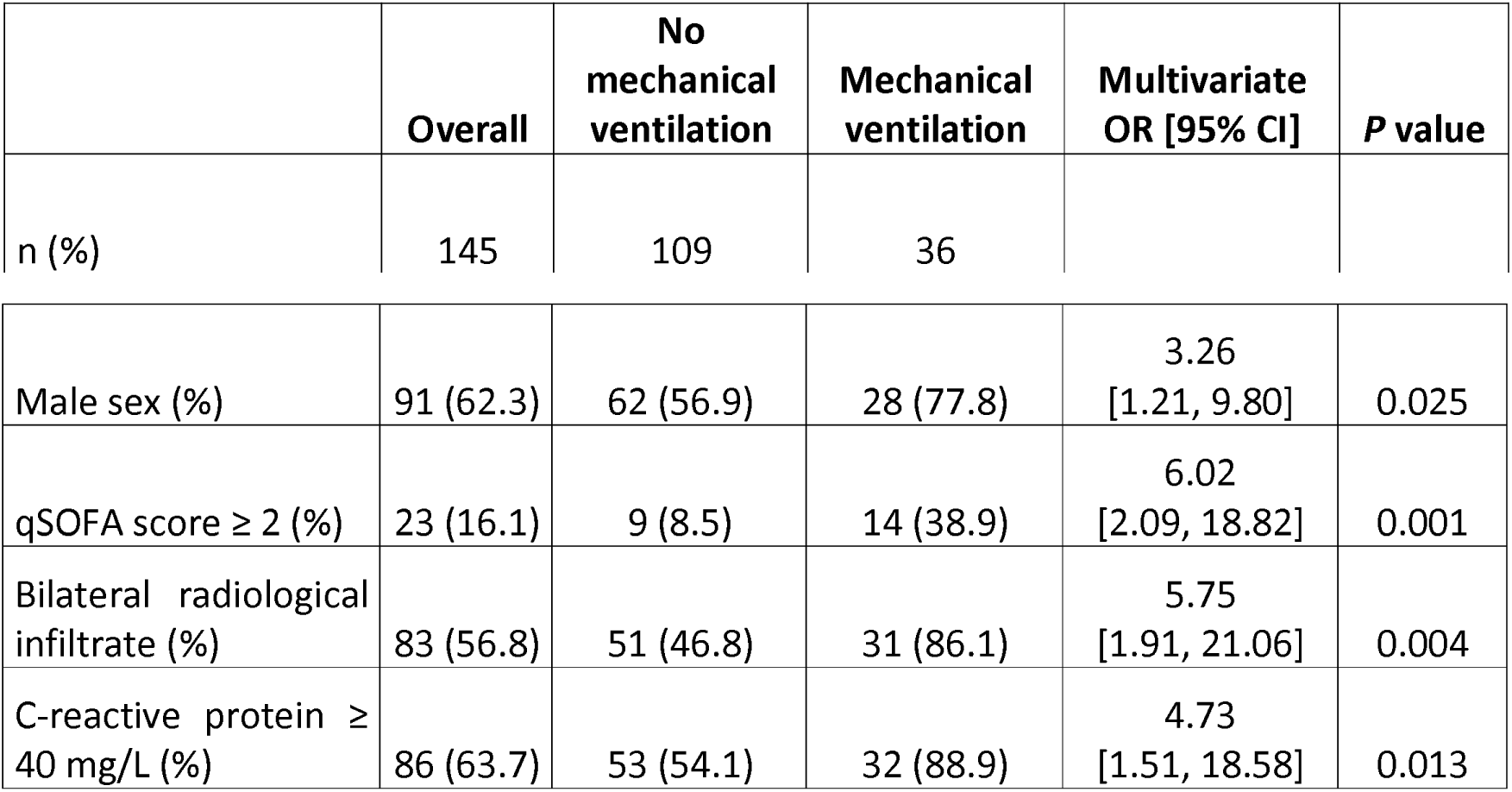
Adjusted risk factors associated with mechanical ventilation at 14 days

## Discussion

Our study identified several risk factors for unfavourable disease progression leading to MV in patients admitted with COVID-19 in a Swiss university hospital.

A quarter of patients for which there were no limitation of care eventually required MV. MV occurred early during the course of hospitalization and the median duration of MV was shorter than previously reported [18]. This effect could result from selection bias of patients with no limitations of care and the limited duration of follow-up.

As infection with SARS-CoV-2 may cause an excessive host immune response, leading to ARDS and death [19]. We would expect biomarkers of inflammation to be associated with unfavourable outcomes. In this study, CRP >40 mg/L on admission was associated with higher odds of MV, suggesting that an unfavourable course is more frequent in patients with a severe inflammatory response related to the infection. Several studies have identified an increased risk of mortality in COVID-19 patients with elevated CRP [17,20]. Other biomarkers (for example, d-dimers) have also been identified as being associated with an increased risk of unfavourable outcome, but we did not identify this link in our study [6]. We believe that CRP is an ubiquitous biomarker whose result could potentially help clinicians assessing the risk of MV in patients with COVID-19. Its use could be easily scaled up due to the availability of numerous point-of-care test.

In our study, we noted an increase in the risk of mechanical ventilation for increasing score values for NEWS, CRB-65 and qSOFA. qSOFA has been proven a useful predictor of mortality among patients with suspected infection [13], mainly of bacterial aetiology [21], but also influenza [22,23]. We opted for including qSOFA in our final multivariate analysis, since it is widely used by clinicians in our institution, and is quick and easy to apply. A higher Sequential Organ Failure Assessment score (SOFA) has been previously linked to increased mortality due to COVID-19 [6]. Data for its calculation are not routinely collected for all patients outside the ICU, making it a less pragmatic tool to quickly evaluate the risk of MV in this patient population.

We additionally identify male sex as a predictor of unfavourable outcome in patients with COVID-19, as previously described [24,25]. It is unclear why there is a sex unbalance in the risk for MV, which could originate in different host response to severe infection [26], notably SARS-CoV-2.

Several factors such as obesity, pregnancy, healthcare worker status, nosocomial acquisition of SARS-CoV-2 infection, previous ACE inhibitors or ARB II treatment and immunosuppressive drugs before admission were not associated with severe disease in this study. Obesity has been described as a risk factor for ICU admission and mortality in patients with influenza [27,28], we found an association between obesity in COVID-19 patients with MV in the univariate, but not the multivariate analysis. Comorbidities generally associated with obesity may contribute in a higher degree to impaired outcomes explaining this finding. We did not identify any relationship between treatment with ECA inhibitors or ARB II and MV, although it has been suggested their use is associated to better outcomes in patients admitted with SARS-CoV-2 [29]. Age was not risk factor for MV: numerous studies have linked age to mortality [2,6], but not to the need for MV. This could be due in part to the limitations of care agreed to in older patients.

More than half of all admitted patients received a treatment with the intent of being specific for SARS-CoV-2, although no evidence for their efficacy exists [30]. A key unanswered question is whether these treatments have an impact on the clinical course of these patients. However, due to indication bias our study cannot answer this question, which should be addressed in prospective randomized clinical trials.

Our study has several limitations to be acknowledged. First, due to its very nature the sample size is limited, which could lead to observation bias in the analyses, with some findings likely to evolve over time. The follow-up period was also limited and several patients were still hospitalised at the time of data analysis. Finally, due to the constantly evolving nature of the epidemic, the clinical care of patients likely evolved during their hospitalisation as changes were made to the recommendations for treatments administered at LUH. The single centre nature of the study limits the generalisability of the results.

However, this study gives some valuable insights in the epidemiology and clinical course of patients admitted in our institution with SARS-CoV-2 infection. We found that male sex, bilateral SARS-CoV-2 pneumonia, elevated CRP and qSOFA equal or greater to two increased the risk of mechanical ventilation. The timely identification of these patients could help us target treatment and better manage the attribution of resources.

## Data Availability

Data is available on Zenodo: https://zenodo.org/record/3799740#.XrkAN2gzaUk

**Abbreviations**
CCI: Charlson Comorbidity Index
AKI: Acute Kidney Injury
ARDS: Acute Respiratory Distress Syndrome
LUH: Lausanne University Hospital
EHR: Electronic Health Records
CRP: C-Reactive Protein
PCT: Procalcitonin
MV: Mechanical ventilation
SOFA score: Sequential Organ Failure Assessment score
qSOFA score: quick SOFA score
NEWS: National Early Warning score
CRB-65 score: Confusion/Respiratory rate/Blood pressure/age≥65 years score
WHO: World Health Organization

## Notes

### Contributors

JR wrote the protocol, participated in data acquisition and validation, performed the data analysis and drafted the manuscript. MP participated in data acquisition and validation, and in drafting the manuscript. RB participated in data acquisition and in drafting the manuscript. PF, JT, FD, BV, EK, LR, DH, MB, CF, KJ and AK participated in data acquisition, PAB, JLP and OM revised the protocol and the manuscript. LS revised the manuscript. LL took part in writing the protocol, in analysing the data, and in drafting the manuscript. All authors contributed to manuscript revision, read and approved the submitted version.

## Supporting information captions

SI Table. Unadjusted risk factors associated with mechanical ventilation at 14 days

## Notes

### Competing Interest Statement

The authors have declared no competing interest.

### Funding Statement

No additional funding was sought for this study.

## References

1 World Health Organization: Novel Coronavirus (2019-nCov) Situation Report-22. World Health Organization, 2020, February 11, 2020. Available: https://www.who.int/docs/default-source/coronaviruse/situation-reports/20200211-sitrep-22-ncov.pdf?sfvrsn=fb6d49b1_2

2. Yang X, Yu Y, Xu J, Shu H, Xia J, Liu H, et al. Clinical course and outcomes of critically ill patients with SARS-CoV-2 pneumonia in Wuhan, China: a single-centered, retrospective, observational study. Lancet Respir Med. 2020;0. doi:10.1016/S2213-2600(20)30079-5

3. World Health Organization: Novel Coronavirus (2019-nCov) Situation Report-22. World Health Organization, 2020, March 11, 2020. Available: https://www.who.int/docs/default-source/coronaviruse/situation-reports/20200311-sitrep-51-covid-19.pdf?sfvrsn=1ba62e57_10

4. https://www.bag.admin.ch/dam/bag/fr/dokumente/mt/k-und-i/aktuelle-ausbrueche-pandemien/2019-nCoV/covid-19-lagebericht.pdf.download.pdf/COVID-19_Situation_epidemiologique_en_Suisse.pdf, accessed on the 1st April 2020. 2020.

5. Bai T, Tu S, Wei Y, Xiao L, Jin Y, Zhang L, et al. Clinical and Laboratory Factors Predicting the Prognosis of Patients with COVID-19: An Analysis of 127 Patients in Wuhan, China. Rochester, NY: Social Science Research Network; 2020 Feb. Report No.: ID 3546118. i Available: https://papers.ssrn.com/abstract=3546118

6. Zhou F, Yu T, Du R, Fan G, Liu Y, Liu Z, et al. Clinical course and risk factors for mortality of adult inpatients with COVID-19 in Wuhan, China: a retrospective cohort study. The Lancet. 2020;0. doi:10.1016/S0140-6736(20)30566-3

7. Grasselli G, Zangrillo A, Zanella A, Antonelli M, Cabrini L, Castelli A, et al. Baseline Characteristics and Outcomes of 1591 Patients Infected With SARS-CoV-2 Admitted to ICUs of the Lombardy Region, Italy. JAMA. 2020 [cited 9 Apr 2020]. doi:10.1001/jama.2020.5394

8. Wynants L, Calster BV, Bonten MMJ, Collins GS, Debray TPA, Vos MD, et al. Prediction models for diagnosis and prognosis of covid-19 infection: systematic review and critical appraisal. BMJ. 2020;369. doi:10.1136/bmj.m1328

9. Corman VM, Landt O, Kaiser M, Molenkamp R, Meijer A, Chu DK, et al. Detection of 2019 novel coronavirus (2019-nCoV) by real-time RT-PCR. Eurosurveillance. 2020;25: 2000045. doi:10.2807/1560-7917.ES.2020.25.3.2000045

10. Harris PA, Taylor R, Minor BL, Elliott V, Fernandez M, O’Neal L, et al. The REDCap consortium: Building an international community of software platform partners. J Biomed Inform. 2019;95: 103208. doi:10.1016/j.jbi.2019.103208

11. Acute Respiratory Distress Syndrome: The Berlin Definition. JAMA. 2012;307: 2526–2533. doi: 10.1001/jama.2012.5669

12. AKI Definition. Kidney Int Suppl. 2012;2: 19–36. doi:10.1038/kisup.2011.32

13. Singer M, Deutschman CS, Seymour CW, Shankar-Hari M, Annane D, Bauer M, et al. The Third International Consensus Definitions for Sepsis and Septic Shock (Sepsis-3). JAMA. 2016;315: 801–810. doi:10.1001/jama.2016.0287

14. Bauer TT, Ewig S, Marre R, Suttorp N, Welte T, CAPNETZ Study Group. CRB-65 predicts death from community-acquired pneumonia. J Intern Med. 2006;260: 93–101. doi:10.1111/j.1365-2796.2006.01657.x

15. McGinley A, Pearse RM. A national early warning score for acutely ill patients. BMJ. 2012;345. doi:10.1136/bmj.e5310

16. Yan L, Zhang H-T, Goncalves J, Xiao Y, Wang M, Guo Y, et al. A machine learning-based model for survival prediction in patients with severe COVID-19 infection. *medRxiv*. 2020; 2020.02.27.20028027. doi:10.1101/2020.02.27.20028027

17. Gong J, Ou J, Qiu X, Jie Y, Chen Y, Yuan L, et al. A Tool to Early Predict Severe 2019-Novel Coronavirus Pneumonia (COVID-19): A Multicenter Study using the Risk Nomogram in Wuhan and Guangdong, China. *medRxiv*. 2020; 2020.03.17.20037515. doi:10.1101/2020.03.17.20037515

18. Ling L, So C, Shum HP, Chan PKS, Lai CKC, Kandamby DH, et al. Critically ill patients with COVID-19 in Hong Kong: a multicentre retrospective observational cohort study. Crit Care Resusc J Australas Acad Crit Care Med. 2020.

19. Xu Z, Shi L, Wang Y, Zhang J, Huang L, Zhang C, et al. Pathological findings of COVID-19 associated with acute respiratory distress syndrome. Lancet Respir Med. 2020;8: 420–422. doi:10.1016/S2213-2600(20)30076-X

20. Lu J, Hu S, Fan R, Liu Z, Yin X, Wang Q, et al. ACP Risk Grade: A Simple Mortality Index for Patients with Confirmed or Suspected Severe Acute Respiratory Syndrome Coronavirus 2 Disease (COVID-19) During the Early Stage of Outbreak in Wuhan, China. Rochester, NY: Social Science Research Network; 2020 Feb. Report No.: ID 3543603. Available: https://papers.ssrn.com/abstract=3543603

21. Maitra S, Som A, Bhattacharjee S. Accuracy of quick Sequential Organ Failure Assessment (qSOFA) score and systemic inflammatory response syndrome (SIRS) criteria for predicting mortality in hospitalized patients with suspected infection: a meta-analysis of observational studies. Clin Microbiol Infect Off Publ Eur Soc Clin Microbiol Infect Dis. 2018;24: 1123–1129. doi:10.1016/j.cmi.2018.03.032

22. Chang S-H, Yeh C-C, Chen Y-A, Hsu C-C, Chen J-H, Chen W-L, et al. Quick-SOFA score to predict mortality among geriatric patients with influenza in the emergency department. Medicine (Baltimore). 2019;98: e15966. doi:10.1097/MD.0000000000015966

23. Papadimitriou-Olivgeris M, Gkikopoulos N, Wüst M, Ballif A, Simonin V, Maulini M, et al. Predictors of mortality of influenza virus infections in a Swiss Hospital during four influenza seasons: Role of quick sequential organ failure assessment. Eur J Intern Med. 2020;74: 86–91. doi:10.1016/j.ejim.2019.12.022

24. Shi Y, Yu X, Zhao H, Wang H, Zhao R, Sheng J. Host susceptibility to severe COVID-19 and establishment of a host risk score: findings of 487 cases outside Wuhan. Crit Care. 2020;24: 108. doi:10.1186/s13054-020-2833-7

25. Du Y, Tu L, Zhu P, Mu M, Wang R, Yang P, et al. Clinical Features of 85 Fatal Cases of COVID-19 from Wuhan: A Retrospective Observational Study. Am J Respir Crit Care Med. 2020 [cited 9 Apr 2020]. doi:10.1164/rccm.202003-05430C

26. Fish EN. The X-files in immunity: sex-based differences predispose immune responses. Nat Rev Immunol. 2008;8: 737–744. doi:10.1038/nri2394

27. Fezeu L, Julia C, Henegar A, Bitu J, Hu FB, Grobbee DE, et al. Obesity is associated with higher risk of intensive care unit admission and death in influenza A (H1N1) patients: a systematic review and meta-analysis. Obes Rev Off J Int Assoc Study Obes. 2011;12: 653–659. doi:10.1111/j.1467-789X.2011.00864.x

28. Sun Y, Wang Q, Yang G, Lin C, Zhang Y, Yang P. Weight and prognosis for influenza A(H1N1)pdm09 infection during the pandemic period between 2009 and 2011: a systematic review of observational studies with meta-analysis. Infect Dis Lond Engl. 2016;48: 813–822. doi:10.1080/23744235.2016.1201721

29. Yang Guang, Tan Zihu, Zhou Ling, Yang Min, Peng Lang, Liu Jinjin, et al. Effects Of ARBs And ACEIs On Virus Infection, Inflammatory Status And Clinical Outcomes In COVID-19 Patients With Hypertension: A Single Center Retrospective Study. Hypertension. 0. doi:10.1161/HYPERTENSIONAHA.120.15143

30. Cao B, Wang Y, Wen D, Liu W, Wang J, Fan G, et al. A Trial of Lopinavir-Ritonavir in Adults Hospitalized with Severe Covid-19. N Engl J Med. 2020. doi:10.1056/NEJMoa2001282

